# Knowledge, Attitudes, and Practices of People living with SCI towards COVID-19 and their Psychological State during In-patient Rehabilitation in Bangladesh

**DOI:** 10.1101/2020.12.21.20248686

**Authors:** Mohammad Anwar Hossain, K M Amran Hossain, Mohamed Sakel, Md. Feroz Kabir, Karen Saunders, Rafey Faruqui, Mohammad Sohrab Hossain, Zakir Uddin, Manzur Kader, Lori Maria Walton, Md. Obaidul Haque, Rubayet Shafin, Iqbal Kabir Jahid

**Affiliations:** Department of Physiotherapy, Centre for the Rehabilitation of the Paralysed (CRP), Jashore, Bangladesh; Department of Microbiology, Jashore University of Science & Technology (JUST), Jashore, Bangladesh; Department of Physiotherapy, Bangladesh Health professions Institute (BHPI), Dhaka, Bangladesh; East Kent Hospitals University NHS Foundation Trust, Canterbury, UK; Department of Physiotherapy & Rehabilitation, Jashore University of Science & Technology (JUST), Jashore, Bangladesh; Kent & Medway NHS and Social care Partnership Trust & University of Kent, Canterbury, UK; John Walsh Centre for Rehabilitation Research, the University of Sydney, Australia and Centre for the Rehabilitation of the Paralysed (CRP), Bangladesh; School of Physical and Occupational Therapy, Faculty of Medicine, McGill University, Montréal, Canada; Institute of Environmental Medicine, Karolinska Institutet, Stockholm, Sweden; Department of Physical Therapy, School of Health Sciences, University of Scranton, Pennsylvania, USA; Department of Physiotherapy, Bangladesh Health Professions Institute (BHPI), Dhaka, Bangladesh; Department of Physiotherapy, Centre for the Rehabilitation of the Paralysed (CRP), Dhaka, Bangladesh

**Author notes:** **Correspondence:** Department of Microbiology, Faculty of Biological Sciences and Technology, Jashore University of Science and Technology, Jashore, 7408, Bangladesh.

## Abstract

**Study Design:** A prospective cross-sectional survey.

**Objective:** The study aimed to examine the Knowledge, Attitudes, and Practices (KAP) of people living with Spinal cord injury (SCI) towards COVID-19 and their psychological status during in-patient rehabilitation in Bangladesh.

**Setting:** The Centre for the Rehabilitation of the Paralyzed (CRP) and the National Institute of Traumatology and Orthopedic Rehabilitation (NITOR), two tertiary level hospitals in Dhaka, Bangladesh.

**Methods:** From July to September 2020, a prospective, cross-sectional survey of SCI subjects, 13-78 years of age, carried out in two SCI rehab centers in Bangladesh. Data has been collected by face to face interview through a pretested, and language validated questionnaire on KAP and Depression, Anxiety, Stress (DASS). Ethical approval and trial registration obtained prospectively. As all the patients were previously living with Spinal cord injury (SCI), therefore, all the patients admitted/ attend SCI rehab centers were considered as SCI positive samples.

**Results:** A total of 207 people with SCI responded, 87%were male, and 13% were female with mean age34.18±12.9 years. 33.8% was tetraplegic and 66.2% was paraplegic and 63.8% of them were diagnosed ASIA-A, with motor score 45.38±19.5, sensory score 97.2±52, SpO2 95.07±3.3, and Vo2max 35.7±3.7mL/kg/min. 178 people had at least one health issue. Overall knowledge score was 8.59±2.3 out of 12, depression 11.18±8, anxiety 7.72±5.1, and stress was 9.32±6.7 from a total of 21 scores each. There was a correlation between Knowledge and DASS with age (P<.05); and Knowledge with gender (P<.05), and education (P<.01). Binary logistic regression found a higher association of Knowledge and DASS with gender (*OR* 6.6, 6.6, .95, 6.6; P<.01); and young age (*OR*.418, P<.01), illiterate (*OR*3.81, P<.01), and rural people (*OR*.48, P<.05) with knowledge. A linear relation was noted between depression and anxiety scores (*r*.45, P<.01) and stress scores (*r*.58, P<.01). A positive attitude was reported for the majority of subjects. SCI Persons reported they and the caregiver followed health advisory in consulting health professionals (65.7%), isolation (63.8%), droplet precaution (87.4%), and hygiene (90.3%).

**Conclusions:** During in-patient rehabilitation in Bangladesh, the majority of SCI reported that they had communicated with health professionals and practiced behaviors that would reduce transmission and risk of COVID-19.

## Introduction

Bangladesh is in a steady status of COVID-19 cases and is expected to experience the second phase of transmission from December 2020 [1]. Institute of Epidemiology, Disease Control and Research (IEDCR) reports 417,475 cases have been diagnosed for COVID-19, among them, 6036 cases died from March 8 2020 to November 8, 2020 [2]. Among the affected cases 54% were aged 21-40 years, males were 71% and among the death cases 39% were aged more than 60, and 29.6% were aged 50-60 years. A low death rate has been observed in active age 21-40 years (11.33%) and a high death rate for Male (77%) among the confirmed cases in Bangladesh [3]. UN called for a disability-inclusive response for COVID-19 [4]. Bangladesh has 10-15% of persons with disabilities (PWD’s) and 80% of them live in rural areas [5-6]. There are many confounding factors and doubts regarding the testing and protection of PWD’s from COVID-19 in Bangladesh [7]. WHO expressed concerns regarding the accessibility of proper information and knowledge on COVID-19 for the PWD’s and urged them to consider the physical, environmental and psychosocial barriers to following the health advisory for them [8].

Sound knowledge on COVID-19 has a relationship with a positive attitude and practicing recommended health advisory in Bangladesh [9]. The study also reflects that using a mask is a key practice along with other advisories to prevent COVID-19 [10]. The study found a strong linear correlation between the knowledge of COVID-19 and the practice of wearing a mask in Bangladeshi general people [9]. Hence, there is no study on Knowledge, attitude and practice towards COVID-19 for any specific group of Persons with disabilities or vulnerable communities in Bangladesh. COVID-19 also has a profound Psychological impact on general people, Bangladeshi people are in fear [9], depressed and anxious [11] and the collective psychological issues may have a long term impact on them [12]. There are profound Psycho-social and economic impacts on persons with disabilities also [13]. Spinal cord injury is a devastating [23, 25], challenging [18], longlasting diverse type of functional impairments that leads to temporary or permanent functional impairments [17]. The survivors of Spinal Cord Injury (SCI) are among the vulnerable communities who require life-long care and face several Physical [14], environmental [15], and psycho-social barriers [16] in the community. Bangladesh has more male dominant, active aged (21-40 years) and complete SCI people who are largely dependent on caregivers and are at risk of several comorbidities [14]. The study recommends SCI people have a real concern of fatality if they are infected with COVID-19 so there is an urgency to protect them [17]. As Bangladeshis have been shown a positive relationship among knowledge, attitude, and practice that is key to prevent COVID-19; there are recommendations from scholarly paper to evaluate the KAP and Psychological status for the people with disabilities also [9].

In the pandemic period, this was not possible to collect data on KAP and the psychological status of SCI people in the community. This study aims to find out the Knowledge, attitude, and practice (KAP) of people living with SCI towards COVID-19 and explore their anxiety, depression, and stress in the pandemic situation during in-patient rehabilitation in Bangladesh.

## Methods

### Study design

The study was a prospective cross-sectional survey of the Persons living with Spinal Cord Injury (SCI) during in-patient rehabilitation in Bangladesh.

### Participants

From July to September 2020 SCI admitted in-patient subjects, all ages, and all genders carried out in two SCI rehab centers in Bangladesh. The patients who are unwilling to respond were excluded from the study. The sample size was not calculated as all the admitted cases from July to September 2020 have been taken as the sample.

### Setting

The study was carried out in the Spinal Cord Injury Unit of the Centre for the Rehabilitation of the Paralyzed (CRP) and the National Institute of Traumatology and Orthopedic Rehabilitation (NITOR).

### Questionnaire

The questionnaire was structured, validated, translated, and customized to fulfill the objectives of the study. The socio-demographic questions (Table 1) were customized to Spinal Cord Injury. The knowledge question was a 12 score pretested questionnaire taken from a Chinese study [18], customized, translated, and used in a population-based study in Bangladesh [9,10]. The health status and co-morbidity questionnaire were modified for SCI respondents and taken from a population-based study [10]. There were Categorical answers to attitudes and beliefs towards the control of the pandemic, and practices of wearing masks and avoiding public gatherings. The Depression, Anxiety, and Stress have been assessed by DASS-21, which is validated [19] to use in SCI subjects. DASS-21 has 21 scores of Depression, Anxiety, and Stress each. For language validation, the questionnaire was translated in forwarding and backward translation to English and Bangla by a group led by a Bilingual British researcher. Later it was evaluated and commented on by a bilingual epidemiologist, a psychiatrist and a clinician experienced working for SCI-related researches.

**Table 1:** Relationship of demographic characteristics with Knowledge and DAS.

| Characteristics |  | Number of participants, n | Knowledge mean± SD | t/Chi | Depression mean± SD | t/Chi | Anxiety mean± SD | t/Chi | Stress mean± SD | t/Chi |
| --- | --- | --- | --- | --- | --- | --- | --- | --- | --- | --- |
| Gender | Male | 180 | 8.46±2.3 | 2.06<sup>a</sup> * | 11.04±7.8 | .669<sup>a</sup> | 7.56±4.9 | 1.174<sup>a</sup> | 9.12±5.8 | 1.108<sup>a</sup> |
|  | Female | 27 | 9.44±1.6 |  | 12.15±9.4 |  | 8.81±6.6 |  | 10.67±11.2 |  |
| Age | 18 years and less | 12 | 8±2.56 | 56.186<sup>b*</sup> | 8.8±6.18 | 82.31<sup>b*</sup> | 6.17±3.9 | 66.884<sup>b*</sup> | 7.50±4.1 | 74.846<sup>b*</sup> |
|  | 19- 40 years | 146 | 9.06±2.07 |  | 11.25±8.1 |  | 7.78±5.35 |  | 9.17±7.1 |  |
|  | 41-60 years | 44 | 7.27±2.67 |  | 11.32±8.2 |  | 8.41±4.9 |  | 10.23±6 |  |
|  | >60 years | 5 | 7.80±1.30 |  | 13.80±7.8 |  | 3.80±2.4 |  | 10.20±7 |  |
| Education | Illiterate | 43 | 8.21±2.0 | 75.22<sup>b**</sup> | 11.26±8.3 | 60.38<sup>b</sup> | 7.95±5.2 | 53.13<sup>b</sup> | 8.70±5.7 | 84.75<sup>b</sup> |
|  | primary education | 54 | 7.87±2.4 |  | 10.28±6.3 |  | 7.43±4.9 |  | 8.13±4.5 |  |
|  | Secondary school | 56 | 9.04±2.1 |  | 11.86±8.1 |  | 7.61±4.6 |  | 10.09±5.9 |  |
|  | Higher secondary | 26 | 8.12±2.3 |  | 11.31±7.81 |  | 8.46±5.4 |  | 7.69±5.4 |  |
|  | Graduation and above | 28 | 10.1±1.9 |  | 11.36±10.12 |  | 7.50±6.4 |  | 12.57±11.7 |  |
| Treatment center | CRP | 146 | 8.53±2.3 | -.528<sup>a</sup> | 11.71±8.1 | 1.45<sup>a</sup> | 8.12±5.6 | 1.69<sup>a</sup> | 10.07±7.4 | 2.48<sup>a*</sup> |
|  | NITOR | 61 | 8.72±2.3 |  | 9.93±7.2 |  | 6.79±3.5 |  | 7.54±4.1 |  |
| Living area | Rural | 139 | 8.58±2.3 | 33.65<sup>b</sup> | 11.60±7.9 | 48.99<sup>b</sup> | 7.6±4.9 | 38.92<sup>b</sup> | 8.86±5.4 | 44.57<sup>b</sup> |
|  | Semi urban | 35 | 7.69±2.3 |  | 8.06±5.3 |  | 7.03±3.6 |  | 7.6±4.3 |  |
|  | Urban | 33 | 9.58±1.9 |  | 12.76±10 |  | 9±7.2 |  | 13.12±11.1 |  |
| Geography | Dhaka | 90 | 8.27±2.1 | 178<sup>b**</sup> | 9.81±6.6 | 152.2<sup>b</sup> | 7.61±4.4 | 130.5<sup>b*</sup> | 8.68±5.3 | 138.1<sup>b</sup> |
|  | Chittagong | 33 | 9.67±.99 |  | 11.45±8.3 |  | 7.27±5.8 |  | 7.09±3.8 |  |
|  | Rajshahi | 7 | 9.14±4.3 |  | 11.71±11 |  | 7.71±6 |  | 10.57±5.3 |  |
|  | Sylhet | 9 | 9.11±1.2 |  | 10.44±8.8 |  | 7.33±3.4 |  | 8.67±6.7 |  |
|  | Khulna | 30 | 8.59±2.1 |  | 13.17±9.7 |  | 8.76±6.3 |  | 10.48±7.7 |  |
|  | Barisal | 19 | 8±2.8 |  | 10.95±5.9 |  | 7.68±5.5 |  | 9.74±6.2 |  |
|  | Rangpur | 12 | 2.50±2.3 |  | 14.83±11.9 |  | 7±6.8 |  | 15.83±14.9 |  |
|  | Mymensingh | 8 | 6.75±3.5 |  | 13.75±6.7 |  | 8.75±3.9 |  | 10.5±6.2 |  |
| ASIA Diagnosis | ASIA A | 132 | 8.68±2.3 | 56.81<sup>b*</sup> | 11.51±8.3 | 52.56<sup>b</sup> | 7.86±5.4 | 39.94<sup>b</sup> | 9.40±7.3 | 51.51<sup>b</sup> |
|  | ASIA B | 10 | 8.60±2.7 |  | 14.81±8.2 |  | 8.40±6.7 |  | 11.4±7.3 |  |
|  | ASIA C | 45 | 8.40±2 |  | 10.49±7.4 |  | 7.51±4.4 |  | 9.02±5.6 |  |
|  | ASIA D | 22 | 8.40±2.7 |  | 8.80±6.5 |  | 7±4 |  | 8.45±4.8 |  |
| Co-morbidity present | Yes | 178 | 8.56±2.3 | 7.11<sup>b</sup> | 11.34±8 | 12.67<sup>b</sup> | 7.81±5.3 | 13.83<sup>b</sup> | 9.62±7 | 15.31<sup>b</sup> |
|  | No | 29 | 8.76±2.1 |  | 10.21±7.8 |  | 6.69±3.5 |  | 7.52±4.2 |  |

<sup>a</sup>Independent t test; <sup>b</sup> Chi-square test; \* Significant with p<.05; \*\* Significant with p<.01

### Data Collection procedure

From July to September 2020, a group of trained data collectors collected the data by face to face interview in the in-patient SCI unit of Centre for the Rehabilitation of the Paralysed (CRP) and the National Institute of Traumatology &Orthopaedic Rehabilitation (NITOR). All the admitted subjects responded to the survey. The questionnaire was asked by the data collector and answers have been documented by them. A total of 207 cases were admitted in the two separate hospitals and all the data has been collected and found eligible (all age groups, willing to participate and no sign and symptoms of COVID 19)to analyze after data audit. The respondents were representing two of the only designated SCI rehabilitation centers in Bangladesh. So it represents all the in-patient SCI peoples in Bangladesh.

### Statistical Analysis

The knowledge score was calculated out of 12 scores as per the correct answers provided and considered as continuous data. The attitude and practice answers were categorical data and DASS-21 is calculated separately in depression, anxiety, and stress out of 21 scores, those are continuous data. Descriptive statistics employed in socio-demographic data, to measure relationships among continuous and categorical data-independent sample t-test for dichotomous data or Chi-square test for more than 2 categories responded. Further Binary logistic regression has been employed using dichotomous variables as the dependent variable and KAP and DASS 21 as covariates. The Pearson test has been used to measure the relationship between two continuous variables. A Chi-square test has been used to determine the relationship between categorical variables. Data analysis was done using the Statistical Package of Social Sciences SPSS version 20.0. The alpha level of significance was set at P <.05.

## Results

### Socio-demographics

The socio-demographic data of the SCI respondent was shown in Table 1. It has been found that 207 respondents responded, 180 were male and 27 were female. The mean age was 34.18± 12.9 years, Majority of the respondents (70.5%)were aged 19-40 years and 21.25% were aged 41-60 years. The majority of the respondents were with secondary education (27%) and 20% of the respondents were illiterate. 70% of the respondents were from CRP and 67% used to live in rural areas. Respondents represent all the administrative divisions of Bangladesh. The majority of the participants (63%) were diagnosed ASIA A and 178 people (85%) reported to have any one of the SCI related complications or COVID-19 related co-morbidities.

### Health status and comorbidity

Among the respondents 33.8% were tetraplegic and 66.2% were paraplegic. 63.8% of the admitted SCI people had been diagnosed as ASIA A. In the International Standard for Neurological Classification of Spinal Cord Injury (ISNCSCI) the mean motor score was 45.38±19.5 out of 100 and the sensory score mean was 97.2±52 out of 253. Oxygen saturation (SpO2) mean was 95.07±3.3 and the maximum rate of oxygen consumption (Vo2max) found was35.7±3.7 mL/kg/min. 178 people (85%) reported having any one of the SCI related complications or COVID-19 related co-morbidities. From multiple response analysis comorbidities are heart disease 2.5%, Hypertension 5%, existing respiratory disease (mostly COPD) 5.7%, Diabetes 1.7%, ulcer and GIT disease 2.7%, Chronic Kidney disease 1%, Chronic liver disease 0.2%, existing pressure sore 18.8%, depressive symptoms 15.1%, urinary tract infection 5.9% postural hypotension 7.7%, bowel bladder complication 20.3% and spasticity 13.3%

### Knowledge

The overall mean knowledge score was 8.59± 2.3 out of 12. Females had a greater mean (12.15±9.4) than males (11.04±7.8). There was a higher knowledge score in the age group 19-40 years (9.06±2.07) but all age groups had a mean of more than 7. The graduates have more knowledge (10.1±1.9) but the illiterate people also have sufficient knowledge of COVID-19 (8.21±2.0). CRP (8.53±2) and NITOR (8.72±2.3) have similar knowledge scores, also the knowledge found satisfactory in rural and urban dwellers. The SCI originated from the Dhaka division has a knowledge score of 8.27±2.1, hence 12 respondents originated from the Rangpur division had a knowledge score of 2.50±2.3. SCI with all the ASIA diagnostic criteria had similar knowledge scores and people with comorbidity had similar knowledge scores (8.56± 2.3) than without comorbidity (8.76±2.1). Statistical association observe with gender and knowledge (t=2.06, p< .05), age with knowledge (X^2^ = 56.18, p< .05), education and knowledge score (X^2^= 75.2, p< .01) and geography with knowledge score (X^2^= 1.78, p< .01). Details are supplied in table 1.

In table 2, Binary logistic regression found relationship with knowledge score and gender (X^2^=11.07, *OR=* 6.6, p< .01), knowledge score with 19 to 40 years vs others (X^2^=29.9, *OR*= .41, p< .01), illiterate vs literate with knowledge score(X^2^= 16.0, *OR*= 3.8, p<.01) and knowledge score with rural vs urban (X^2^= 11.4, *OR*= .48, P<.01). Table 3 shows, no statistical significant of knowledge score has been found with family income (*r*=.107,P=.126), depression (*r=*-.06, P=.39), anxiety (*r=*.04, P=.49) and stress (*r*= .05, P=.42).

**Table 2.** Results of Binary logistic regression on factors associated with Knowledge and DASS.

| Variables | Knowledge |  |  |  | Depression |  |  |  | Anxiety |  |  |  | Stress |  |  |  |
| --- | --- | --- | --- | --- | --- | --- | --- | --- | --- | --- | --- | --- | --- | --- | --- | --- |
|  | Chi | Coefficient | OR | p | Chi | Coefficient | OR | p | Chi | Coefficient | OR | p | Chi | Coefficient | OR | p |
| Male vs Female | 11.075 | 1.89 | 6.6 | .001** | 28 | 1.89 | 6.6 | .001** | 1.294 | -.043 | .958 | .243 | 1.108 | 1.89 | 6.6 | .001** |
| 19-40 years vs others | 29.97 | -.873 | .418 | .001** | 12.38 | -.873 | .418 | .003** | .059 | -.007 | .993 | .808 | .248 | .011 | 1 | .61 |
| Illiterate vs literate | 16.033 | 1.33 | 3.81 | .001** | 12.33 | 1.33 | 3.8 | .03* | .105 | -.011 | .989 | .744 | .494 | .019 | 1 | .49 |
| CRP vs NITOR | 3.2 | -.873 | .418 | .013* | 16.57 | -.873 | .418 | .04* | 3.02 | -.055 | .947 | .09 | 7.14 | -.072 | .93 | .01* |
| Rural vs Urban | 11.42 | -.715 | .48 | .003** | 17.52 | -.715 | .48 | .04* | .255 | .014 | 1 | .612 | 1.97 | .030 | 1 | .163 |
| Dhaka vs others division | 13.9 | .262 | 1.3 | .06 | 27.78 | .262 | 1.3 | .06 | .077 | .008 | 1 | .782 | 1.50 | .026 | 1 | .23 |
| Complete SCI vs incomplete SCI | 12.45 | -.565 | .568 | .02 | 14.05 | -.565 | .568 | .003** | .238 | -.014 | .986 | .628 | .049 | -.005 | .995 | .826 |

\* Significant with $p < .05$ ; \*\* Significant with $p < .01$ , vs means versus

**Table 3:** Relationship among Family income, knowledge and DASS.

|  | Family income | Knowledge | Depression | Anxiety |
| --- | --- | --- | --- | --- |
| Family income |  |  |  |  |
| Knowledge | $r = .107$<br>P .126<br>N= 207 | | | |
| Depression | $r = -.060$<br>P .390<br>N= 207 | $r = -.049$<br>P .483<br>N= 207 | | |
| Anxiety | $r = .047$<br>P .499<br>N= 207 | $r = -.041$<br>P .562<br>N= 207 | $r = .452^{**}$<br>P .001<br>N= 207 | |
| Stress | $r = .054$<br>P .438<br>N= 207 | $r = .058$<br>P .404<br>N= 207 | $r = .586^{**}$<br>P .001<br>N= 207 | $r = .476^{**}$<br>P .001<br>N= 207 |

\* Significant with $p < .05$ ; \*\* Significant with $p < .01$

### Attitude

The majority of the respondent beyond age, gender, geography had a positive attitude towards preventing COVID-19. Table 4 shows, the statistical relationship between education and belief in control (X^2^= 14.0, p<.05), living area with belief in control (X^2^= 9.4, p< .05), and geography with a positive attitude (X^2^= 4.19, p<.05).

**Table 4:** Relationship among Attitude and practice with demographic variables

| Variables |  | Attitude: Belief in Control |  |  | Attitude: World can win |  |  | Practice: Go to crowd |  |  | Practice: Wear Mask |  |  |
| --- | --- | --- | --- | --- | --- | --- | --- | --- | --- | --- | --- | --- | --- |
|  |  | Agree | Disagree | Chi value | Yes | No | Chi | Yes | No | Chi | Yes | No | Chi |
| Gender | Male | 125 | 55 | 3.29 | 117 | 63 | 1.74 | 58 | 121 | .38 | 146 | 33 | .151 |
|  | Female | 14 | 13 |  | 14 | 13 |  | 11 | 17 |  | 23 | 5 |  |
| Age | 18 years & less | 11 | 1 | 6.93 | 10 | 2 | 3.47 | 4 | 8 | 4.35 | 9 | 3 | 17.6* |
|  | 19- 40 years | 100 | 46 |  | 92 | 54 |  | 50 | 96 |  | 128 | 18 |  |
|  | 41-60 years | 24 | 20 |  | 25 | 19 |  | 14 | 30 |  | 28 | 16 |  |
|  | >60 years | 4 | 1 |  | 4 | 1 |  | 1 | 4 |  | 4 | 1 |  |
| Education | Illiterate | 21 | 22 | 14.08* | 22 | 21 | 4.41 | 11 | 32 | 6.0 | 34 | 9 | 11.04 |
|  | primary | 36 | 18 |  | 35 | 19 |  | 18 | 36 |  | 43 | 11 |  |
|  | Secondary school | 46 | 10 |  | 40 | 16 |  | 24 | 32 |  | 51 | 5 |  |
|  | Higher secondary | 15 | 11 |  | 16 | 10 |  | 8 | 18 |  | 17 | 9 |  |
|  | Graduation/above | 21 | 7 |  | 18 | 10 |  | 8 | 20 |  | 24 | 4 |  |
| Treatment center | CRP | 101 | 45 | .924 | 95 | 51 | .678 | 52 | 94 | 1.45 | 120 | 26 | .507 |
|  | NITOR | 38 | 23 |  | 36 | 25 |  | 17 | 44 |  | 49 | 12 |  |
| Living area | Rural | 84 | 55 | 9.4* | 79 | 60 | 8.84* | 42 | 97 | 6.4 | 117 | 22 | 8.2 |
|  | Semi urban | 30 | 5 |  | 29 | 6 |  | 14 | 21 |  | 25 | 10 |  |
|  | Urban | 25 | 8 |  | 23 | 10 |  | 13 | 20 |  | 27 | 6 |  |
| Geography | Dhaka | 65 | 25 | 1.85 | 64 | 26 | 4.19* | 38 | 52 | 6.4* | 76 | 14 | 2.07 |
|  | Other part | 74 | 43 |  | 67 | 50 |  | 31 | 86 |  | 93 | 24 |  |
|  | ASIA A | 83 | 49 | 4.33 | 80 | 52 | 3.02 | 43 | 89 | 7.5 | 110 | 22 | 17.9* |
|  | ASIA B | 7 | 3 |  | 7 | 3 |  | 5 | 5 |  | 4 | 6 |  |
|  | ASIA C | 32 | 13 |  | 28 | 17 |  | 12 | 33 |  | 36 | 9 |  |
|  | ASIA D | 17 | 3 |  | 16 | 4 |  | 9 | 11 |  | 19 | 1 |  |

\* Significant with $p < .05$ ; \*\* Significant with $p < .01$ ; CRP= Centre for the Rehabilitation of the Paralyzed, NITOR= National Institute of Traumatology & Orthopedic Rehabilitation

### Practice

The majority of the respondents (66%) did not go to the crowd and 80% wear a mask during every exposure. 32% of the male respondents went to the crowd at least one time and 39% of the female respondents went to the crowd once during the lockdown period. 18% of the male did not wear the mask and 17% of females did not wear a mask during exposure to outside people. There was a statistically significant relationship found with age and practice of wearing the mask (X^2^= 17.6, p<.05), geography with staying isolated (X^2^=6.4, p<.05), and complete SCI with practice wearing mask (X^2^=17.9. p,.05). Details are in table 4. SCI Persons reported they and the caregiver followed health advisory in consulting health professionals (65.7%), isolation (63.8%), droplet precaution (87.4%), and hygiene (90.3%).

### Depression

The overall mean depression score was 11.18±8 out of 21. Females had higher depression (12.15±9.4) than men (11.04±7.8). Elderly people, more than 60 years old had higher depression (13.8±7.8) than other age groups. Urban people had more depression (12.76±10) than rural or semi-urban dwellers. The persons with SCI having comorbidities showed a higher level (11.34±8) of depression. There was a relationship between depression with age (X^2^=82.31, p<.05) (Table 1). Table 2 shows, Male had more depression than females (X^2^=28, *OR*=6.6, p<.01) and the young age people aged 19 to 40 years had more depression than others (X^2^=12.38, *OR*=.418, p<.01). illiterate people were in high depression than literate (X^2^=1.33, *OR*=3.8, p<.05), rural people showed higher depression than urban people and SCI people diagnosed with ASIA A were highly depressed than incomplete SCI (X^2^=14.05, *OR*=.56, p<.01). Table 3 represents statistical relationships among depression and anxiety (*r*= .45, p<.01), and depression with stress (*r*= .58, p<.01).

### Anxiety

The overall mean depression score was 7.72±5.1 out of 21. Table 1, represents females (8.81±6.6) had a higher anxiety score than males (7.56±4.9). Persons aged 41 to 60 years had more anxiety (8.41±4.9), the person having higher secondary education (8.46±5.4), urban people (9±7.8), incomplete paraplegic (8.40±6.7), and the person with comorbidities shown higher anxiety level (7.81±5.3). Binary logistic regression did not find any relationship between the anxiety score and any specific group variables. Pearson correlation found highly statistical significance (p<.01) of the relationship between anxiety with depression (*r*= .45) and anxiety with stress (*r*=.47) (table 3).

### Stress

The overall mean depression score was 9.32± 6.7 out of 21. Females had higher stress (10.67±11.2) than males (9.12±5.8) (table 1). Binary logistic regression results similar (X^2^=1.108, OR= 6.6, p<.01) (Table 2). In table 3,the relationship was found with stress and depression (*r*= .58, p<.01) and anxiety (*r*=.476, p<.01) (Table 3).

## Discussion

The survey aimed to find out the Knowledge, attitude, and practice (KAP) of people living with SCI towards COVID-19 and determine their anxiety, depression, and stress during in-patient rehabilitation in Bangladesh. All of the 207 in-patient rehab patients admitted in two dedicated spinal injury units of two renounced rehabilitation centers from July-September 2020 responded to the survey. Data are scarce on the exact prevalence of SCI; hence CRP accommodates 390 SCI patients that is one of the largest facilities in LMIC’s of the World [20]. SCI’s are one of the most vulnerable groups of persons with disabilities having diverse Physical, environmental and psycho-social impairments [20] and perceived death of every one of five people within 2 years of discharge from hospital [21]. The survey found a satisfactory overall knowledge about COVID-19 (8.59±2.3 out of 12), and a positive attitude and practice with the general population (8.71±1.64 out of 12) in Bangladesh [9]. The reason might be a series of patient education and awareness activities [22] made by the rehabilitation centers during their uninterrupted service provision in the COVID-19 Pandemic. There were nearly 36 COVID-19 positive cases among the respondents who were later diagnosed by Real-Time Polymerase Chain Reaction (RT-PCR) test at CRP, a few of them needed critical care but none reported the death.

The materials and tools were satisfactory. Knowledge, Attitude, and Practice (KAP) questions were validated and used in a population-based survey of 2157 respondents in Bangladesh [10]. The knowledge score was more in females in this study, similar findings attributed in that population-based survey. Additionally, some questions were added related to the practice of additional health advisory for the SCI people and their caregivers. DASS-21 is validated for SCI cases [19], hence the appropriate process of forwarding and backward translation and expert opinion has been taken before using the tools.

The respondents were male-dominant aged 19-40 years, the majority was diagnosed as ASIA-A, were rural dwellers, and with a co-morbidity. This is a common feature as young males living in rural areas are more prone to risks associated with SCI, and five years epidemiological study reports more than 80% of admitted SCI’s are male, 55% with age range 19-40 years, 69% lived in rural areas and 60% were diagnosed as ASIA-A [23]. The co-morbidity of SCI people is common in many aspects ranging from pain, respiratory issues, pressure ulcers, cardiovascular, nephrology, and physical inactivity in almost every SCI survivors; the ambulatory people have less complication than bed-ridden people [20-21]. The respondents were admitted to hospital and mostly bed-ridden or dependent on a caregiver, these cumulative factors made an SCI survivor one of the vulnerable groups to the fatality of COVID-19 in Bangladesh.

SCI people and their caregivers had a positive attitude to the control of COVID-19 and they mostly followed (>80%) the health advisory during in-patient rehabilitation. The reason behind the findings might be the successful patient protective policy of CRP and NITOR during the COVID-19 pandemic, moreover, the similar positive attitude and practice noted in a population-based study in Bangladesh [10].

The overall depression score 11.18±8, anxiety 7.72±5.1, and stress 9.32±6.7 from a total of 21 scores each seems different from the population assumption. The depression level was a little higher as 15% of the respondents had existing issues with depression. The majority of psychological issues were relevant to the male gender, young age, and complete patients. In a web-based survey majority of the male university students had mild depressive symptoms in Patient Health Questionnaire [24]. Another Bangladeshi study [20] also supports having a lower depression level in the majority of SCI people in Bangladesh. Hence anxiety and stress are interrelated and a linear relation was noted between depression and anxiety scores (*r*.45, P<.01) and stress scores (*r*.58, P<.01). A population-based study reveals, nearly 86% of Bangladeshi people have stress issues related to COVID-19 [25]. The anxiety and stress score may be associated with the affordability, safety, and reliability issues of the rehabilitation centers that were not examined in this survey.

There was a correlation between Knowledge and DASS with age (P<.05); and Knowledge with gender (P<.05), and education (P<.01). Binary logistic regression found a higher association of Knowledge and DASS with gender (P<.01); and young age (P<.01), illiterate (P<.01), and rural people (P<.05) with knowledge. The findings are identical to the similar population-based study [9].

The study limits the SCI survivors during in-patient rehabilitation. In this period they are cared for by the health professionals and surrounded by the protection of an institution based Rehabilitation. WHO reported non-communicable disease services and rehabilitation to be mostly interrupted or affected services during the COVID-19 pandemic worldwide [12]. The study reflects a positive role of institutions to take necessary steps on the protection, health promotion, and rehabilitation of the SCI’s in the COVID-19 pandemic in Bangladesh.

There are concerns about the safety and protection of People with SCI living in the community [26]. Future studies of KAP, psychological aspects, coping on COVID-19 and the livelihood issues of the persons living in the community with Spinal Cord Injury are recommended.

## Conclusion

During in-patient rehabilitation in Bangladesh, the majority of SCI reported that they had communicated with health professionals and practiced behaviors that would reduce transmission and risk of COVID-19. The Institute based rehabilitation to SCI was appropriate to protect these vulnerable SCI people from the COVID-19 catastrophe in Bangladesh.

## Data Availability

The data is achieved in a public repository and accessible

https://www.kaggle.com/kmamranhossain/kap-sci-bd

## Data Archiving

The data is achieved in a public repository and accessible at https://www.kaggle.com/kmamranhossain/kap-sci-bd

## Acknowledgments

The authors acknowledge Professor Dr. Md. Forhad Hossain for statistical analysis and the data collectors.

## Statement of Ethics

The ethical permission obtained from CRP Ethical review committee (CRP-R&E-0401-296) and trial registration obtained prospectively from the primary trial registry site of WHO (CTRI/2020/06/025529). Also, written permission is taken from the authority of both centers. Respondents have given written consent or relatives gave consent on behalf of them. The participation was voluntary.

## Conflict of Interest

The authors declare no competing interest. The research awarded the best oral presentation at the Asian Spinal Cord Network Conference 2020. All the authors participated in the study from their involvement without their institutional interest or support.

## Author contributions

MAH & KMAH: conceptualization, data curation, methodology, formal analysis, and write-up. IKJ & MS: conceiving and designing the study and major revisions. KS, RF, LMW: data curation and overall editing. ZU, MK, and MSH: input on methodology, analysis, results, and discussion section. ER and PL: assisted with the writing of results and overall editing. MFK, MOH & RS: designing the study.

## Funding

The study was self-funded research of the authors.

---

**Figure 1:**
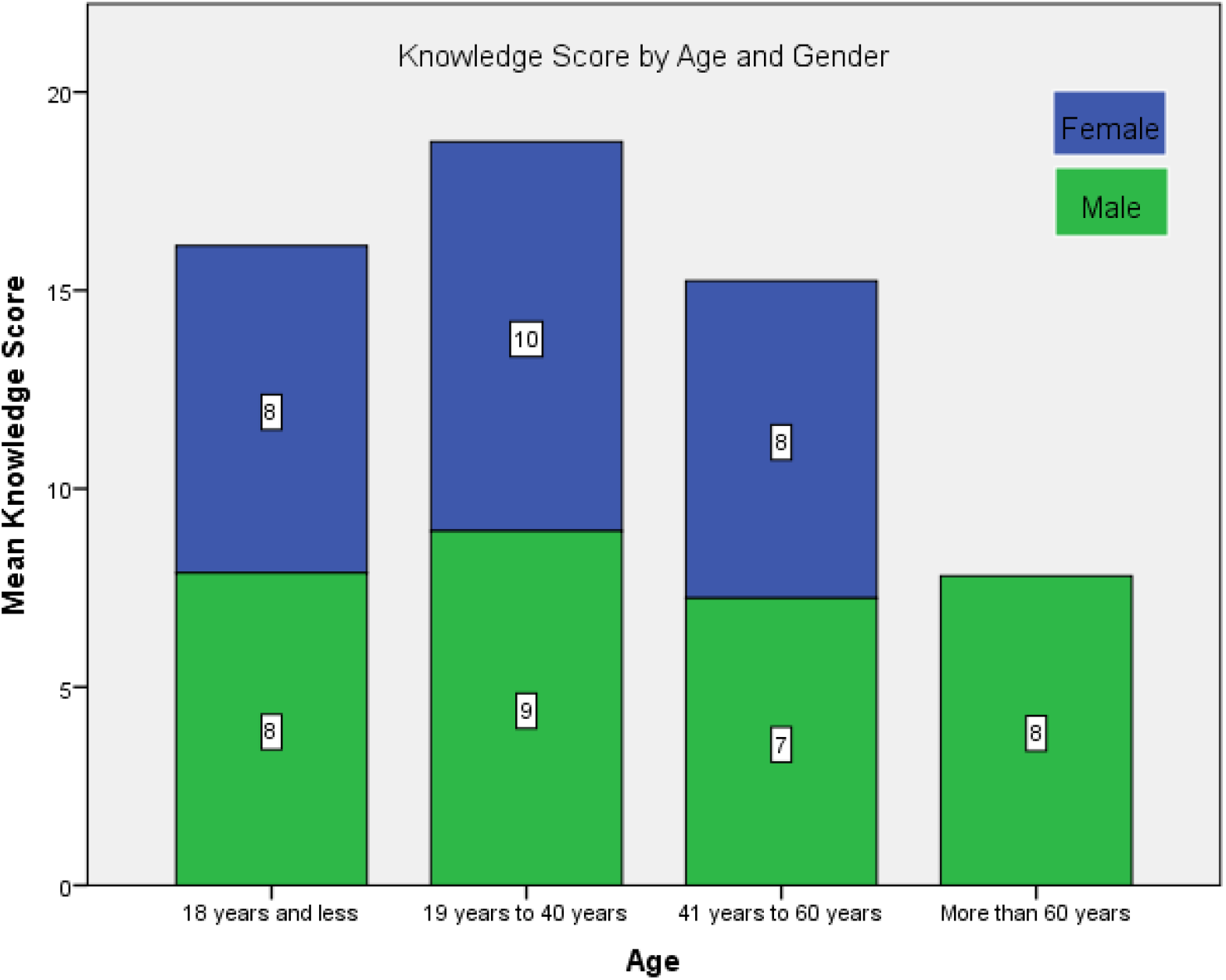
Knowledge score by Age and Gender.

## Notes

### Competing Interest Statement

The authors have declared no competing interest.

### Clinical Trial

Observational study hence trial registration is not required

### Funding Statement

Self-funded study

